# The age-stratified analytical model for the spread of the COVID-19 epidemic

**DOI:** 10.1101/2021.07.13.21260459

**Authors:** F. Mairanowski, Below

## Abstract

The previously developed ASILV model for calculating epidemic spread under conditions of lockdown and mass vaccination was modified to analyse the intensity of COVID-19 infection growth in the allocated age groups.

Comparison of the results of calculations of the epidemic spread, as well as the values of the seven-day incidence values with the corresponding observation data, shows their good correspondence for each of the selected age groups.

The greatest influence on the overall spread of the epidemic is in the 20-40 age groups. The relatively low level of vaccination and the high intensity of contact in these age groups contributes to the emergence of new waves of the epidemic, which is especially active when the virus mutates and the lockdown conditions are relaxed.

The intensity of the epidemic in the 90+ age group has some peculiarities compared to other groups, which may be explained by differences in contact patterns among individuals in this age group compared to others.

Approximate ratios for estimating mortality as a function of the intensity of infection for individual age groups are provided.

The proposed stratified ASILV model by age group will allow more detailed and accurate prediction of the spread of the COVID-19 epidemic, including when new, more transmissible versions of the virus mutate and emerge.

## Introduction

In contrast to numerous models of epidemic spread of COVID-19 infection which rely on numerical methods, the analytical model proposed herein enables us to obtain all relevant characteristics of the epidemic in a timely manner without resorting to computer software development. The model, named ASILV [1, 2, 3, 4, 5, 6, and 7],can be used to determine both the time-varying total number of infected persons and the incidence, i.e. fluctuations in the epidemic level over a certain period of time.

Despite the simplicity of the methodology and the advantages of having to use only two semi-empirical coefficients, the results obtained using the model agree quite satisfactorily with the corresponding observational data. For example, the average correlation between estimated and statistical data on epidemic intensity ranges from 95% to 99%.

The model is based on the well-known but modified system of equations previously used in the SIR model [8]. Under certain assumptions concerning the relationship between individuals infected by I and those susceptible to infection by S, taking into account the impact on epidemic growth of lockdown, which has efficiency L, of mass vaccination intensity V, the analytical dependence has been obtained. In particular, it has been established [6] that the epidemic development is determined by three dimensionless complexes composed of the intensities: transmission, vaccination and restrictions of virus spread by lockdown. The obtained analytical relation makes it possible to analyse the effectiveness of lockdown and assess the possible consequences of its partial or total cancellation at various mass vaccination intensities.

The most important characteristic of an epidemic, which determines the conditions for its development, is the ability of a virus to mutate in such a way that new strains acquire properties substantially different from previous variants. It can be assumed that as the number of infections increases, the likelihood of such mutations will increase. The general trend of changing the properties of the virus is to increase its transmissibility. Consequently, with decreasing lockdown in populations not exposed to mass vaccination the probability of mutation of the virus can increase. Since the general strategy of vaccination is to give priority to the so-called “risk groups” and above all the older population groups, younger segments of the population are less protected against infection. Under these circumstances, the use of a single model for all population groups is not entirely correct. It is necessary to develop a model to calculate the epidemic separately for each of the age groups. This is the task of this paper.

## Methodology

The development of an age-stratified model of epidemic spread is usually based on the so-called contact matrix [9]. A contact matrix consists of a square with the age groups, along the horizontal and vertical sides, divided into which the entire population is divided. The use of the contact matrix makes it possible to determine the closeness of contacts between different age groups and, when performing numerical calculations using a particular model, to establish patterns of epidemic development for each allocated class. Under normal conditions, without the introduction of lockdown, a typical contact matrix is characterized by higher values of contacts along the diagonal of the square, indicating a closer interaction within age groups, e.g. children in schools, teenagers in universities, adults of a certain age at work, elderly people in old people’s homes.

The introduction of lockdown disrupts this established social structure, the closeness of contacts within individual age groups is sharply reduced, and the matrix of contacts becomes much more homogeneous [10, 11, 12, 13]. For example, according to data cited for France, the introduction of lockdown has reduced the number of contacts exactly tenfold, at that, the number of contacts within the population aged under 60 has been approximately twice as high as in the older age-groups. Finally [13], the global matrix depicts a higher density of within age group contacts.

Thus, for simplicity of the problem formulation we will single out separate age groups under lockdown conditions, considering only contacts within each of them. In case of poor agreement between the calculated and observed values it is possible to correct the calculation by changing the model coefficient. The initial equations in this case do not differ from those previously used to calculate the spread of the epidemic for the entire population [6]. The basic formula derived from the solution of these equations for calculating the spread of an epidemic is as follows:

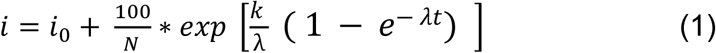

*i*- is the relative number of infected persons per one inhabitant of the settlement in question, as a percentage,

λ - intensity factor of decrease in contacts of infected patients with persons who potentially can get infected by means of quarantine and other preventive measures,

N - total population of the area under consideration,

i_0_ - is the value of I at the initial moment of the calculation period,

k _-_ is the transmission rate coefficient for the settlement with a population of N, which is calculated by the formula [7]:

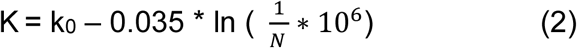

The K coefficient also depends on the transmissibility of the virus strain responsible for the spread of the epidemic during the time period in question. The influence of the transmissibility of the virus is taken into account by changing the parameter k_0_. For the original strain, the coefficient was assumed to be 0.355. For other strains γ times more transmissible than the original strain, the coefficient is determined using a simple formula

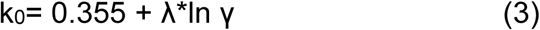

Keeping in mind that λ = 0.034 for the average conditions in the case of predominant action, we find k_0_ = 0.37 for the so-called α strain, whose transmissibility is about 60% higher than the original strain (γ = 1.6), and k_0_ = 0.385 for even more active δ variant strain.

As noted in [6], the development of the epidemic may be influenced by climatic factors. The dependence of the K coefficient on temperature and UV value can be approximated using the parameter

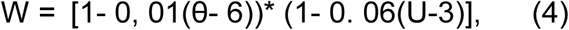

where W is coefficient of influence of climatic parameters on intensity of epidemic development, Δ*i*_*max*.*w*_ – is maximum intensity of infection growth taking into account climatic factors, θ is average air temperature (C^0^), U is value of UV index (for average conditions of Berlin it is assumed that θ = 6 C^0^, U=3). Accordingly, in addition to equation (3), we have:

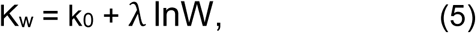

where Kw is the coefficient in equation (2), taking into account the influence of climatic factors on it.

If the spread of infection is associated with several virus strains, the calculated dependence will be written in the following form [6]:

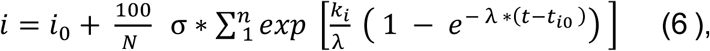

where:

*k*_*i*_- the transmission rate coefficient of the new virus strain and the time of the epidemic wave associated with the new coronavirus strain

*t*_*i*0_ - the start time of the new epidemic wave associated with the new coronavirus strain.

σ - Heaviside symbol σ = 1 Пpи t ≥ *t* _*i*_ и σ = 0 at t < *t*_*i*_

When mass vaccination is carried out, fort ≥ *t* _*ν*_:

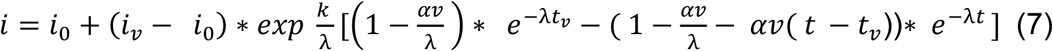

where

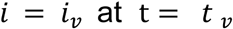

The following additional notations have been introduced into equations and (7):

v - population vaccination rate (1/day)

α - coefficient of vaccine effectiveness,

*t*_*v*_ - time when vaccination starts

The same equation (7) is used to calculate epidemic progression subject to dramatic changes in vaccination rates, as has been done, for example, in many European countries, notably Germany. The model equations presented were originally given in [7].

The model coefficient λ is related to the effectiveness of the lockdown condition. To this end, the parameter L, characterizing the level of reduction in the epidemic growth rate due to lockdown, is introduced [7]:

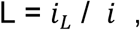

where

*i*_*L*_ - and i are the epidemic growth rate with and without lockdown, respectively.

For example, if application of lockdown reduces the maximum number of infected residents by half, then the coefficient L = ½ = 0.5. Using dependence (1) for time t → ∞, varying the value of coefficient λ, we find the relationship between this coefficient and the parameter L. The graph of this dependence is shown in Fig. 1.

**Figure 1.**
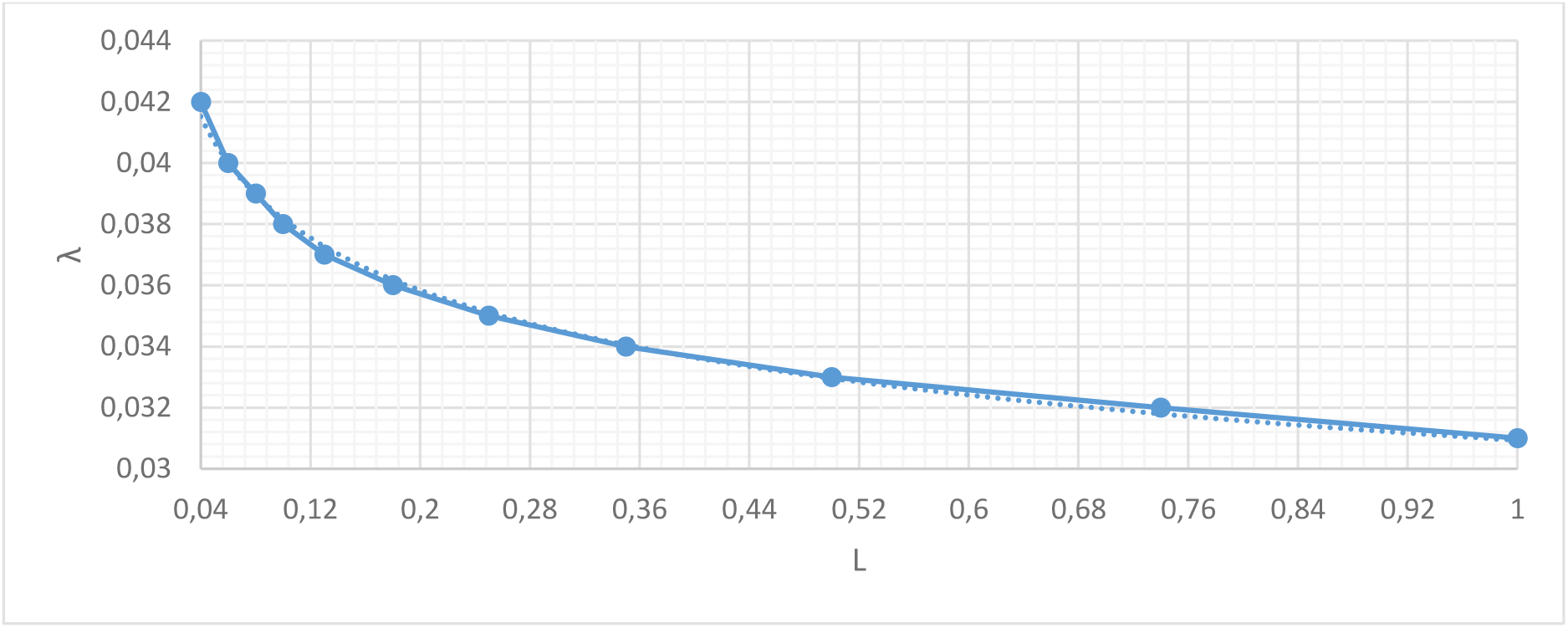
Dependence of the model coefficient λ on the effectiveness of the lockdown L.

This graph shows, in particular, that in the absence of lockdown, the coefficient λ can be assumed to be 0.031/day, and that when this coefficient is above 0.042 1/day, the epidemic wave is virtually suppressed by lockdown. However, this does not exclude the possibility of a new virus strain emerging when the lockdown conditions are relaxed. For the most characteristic lockdown conditions in most European countries, a coefficient of λ = 0.034-0.035 1/day can be assumed; hence, the L coefficient varies between 0.2 and 0.3, which means that lockdown reduces the epidemic’s growth rate by a factor of 3-5.

The proposed calculation methodology can be used separately for each of the allocated age groups.

## Results

We calculate the epidemic spread of the virus for each of the age groups selected from the general population in Berlin. A total of 10 groups are distinguished, from ages 0-9 to 90+. The numbers of each group are determined according to the data given in [14]. The age groups of the standard statistical data do not correspond to the age groups used in the infectiousness analysis. As a consequence of this discrepancy, we had to estimate the size of each age group according to the ratio of the total number of cases per 100,000 inhabitants for each group.

The start of the calculated epidemic wave in Berlin was taken as 04.09.2020, when a monotonic increase in the number of infected persons was recorded [7].

About 140 days after the outbreak of the second wave of the epidemic in Berlin, there were noticeable signs of the emergence of a new so-called “British” or α- strain of the virus. Virological studies have shown that this virus strain has a slightly higher transmissibility than previous strains [15]. In our model this characteristic of the “British” strain was taken into account by increasing the K coefficient according to relation (3) by 0.015. As a consequence of the steep rise in epidemic intensity during this period, additional restrictive lockdown measures were introduced in Berlin. For this period, the tightening of lockdown conditions was taken into account in the calculations by increasing the coefficient λ.

Since the end of January 2021, mass vaccination of the population has been carried out in Berlin. However, the intensity of vaccination in its initial phase was very low [14]. In the first phase it was mainly the elderly over 80 years of age, followed by the 60+ age group and the “risk group”. Young people aged 18 years and over were only vaccinated from the end of May, but we were unable to find specific information on the number of infected persons for each age group in Berlin. In [16], statistical data on the dynamics of vaccination depending on age are given for a number of countries. By analysing these data for several European countries (Austria, Belgium, Denmark, France, Ireland, Italy, Spain, Sweden and Norway), an age-dependent estimate of the vaccinated population in Berlin (as of 25.06.2021) was calculated. For the age group over 60 years old the vaccinated part of the population is 70-90%. For populations aged between 19 and 60, the vaccination rate was assumed to be between 5% and 20%, depending on age. However, vaccination accounting for these populations does not affect the results of the epidemic growth calculations, as the duration of vaccination for them does not exceed one month.

Given the effectiveness of vaccines in preventing transmission, we obtain a dependence for calculating the effective vaccination rate:

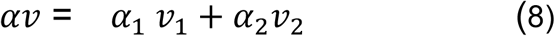

Vaccination rates for each vaccine dose *ν*_1_ and *ν*_2_were calculated as the ratio of the percentage of vaccinated population (for each age group) to the total time of mass vaccination of the population. BioNTech - Pfizer and Moderna vaccine effectiveness ratios for the first and full doses of vaccination were taken to be *α*_1_ = 0.7, *α*_2_ = 0.92, respectively [17]. These effectiveness coefficients are obtained for the so-called alpha variant of the virus. As the virus mutates, more resistant variants of the virus may emerge, which will contribute to a reduction in vaccination effectiveness.

In the following, for simplicity of calculation, the effect of changes in air temperature and ultraviolet radiation on the intensity of infection growth was not taken into account. In addition, changes in lockdown conditions and, consequently, changes in the value of the coefficient λ during the Christmas and Easter holidays were not taken into account. If necessary, these adjustments can be introduced into the calculations, as in our previous studies [6, 7].

The results of calculations according to the proposed methodology for age groups 0-9 and 10-19 years old are shown in Figure 2. The same graph shows the observation data for both the corresponding age groups and for Berlin as a whole [14].

**Fig. 2.**
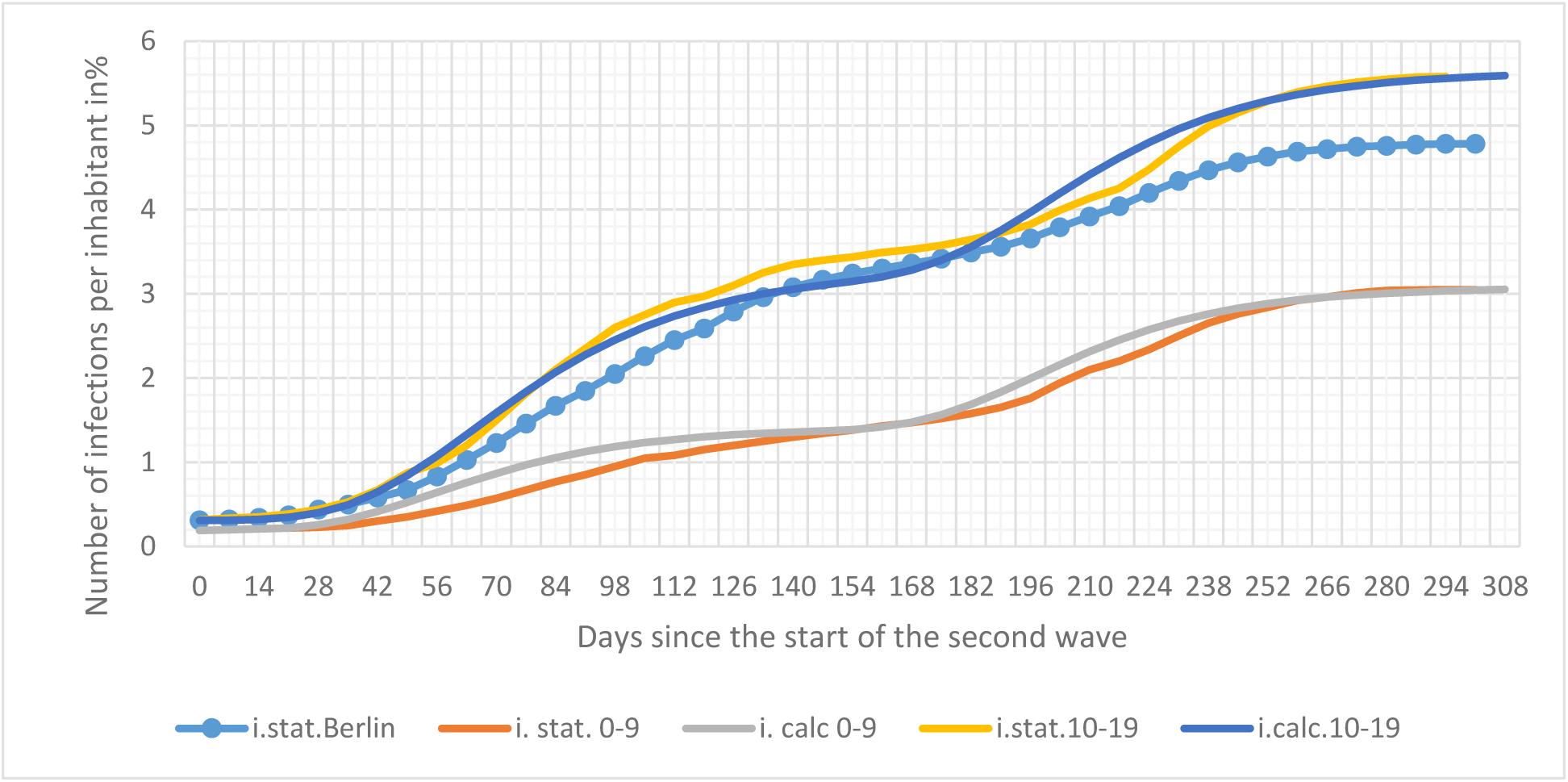
Spread of the epidemic in the age groups 0-9 and 10-19 (i.calc. 0-9, i.calc. 10-19 – calculated infections for age groups 0-9 and 10-19 years respectively, i.stat.Berlin, i.stat.0-9, i.stat.10-19 - observed data of infections for Berlin and for age groups 0-9 and 10-19 years respectively)

Calculations for the 0-9-year-old group were performed for a value of λ = 0.038 1/day. Such a high value of the coefficient λ for this age group can be explained by the high degree of isolation of children of this age group during the lockdown period, both from their peers and from other people. For the 10- to 19-year-old age group, the intensity of the epidemic growth rate appears close to the Berlin average, with a coefficient λ = 0.034 1/day for the initial stage and a narrowing of the lockdown conditions, starting at time t = 140 days, this coefficient increases to λ = 0.036 1/day.

In the analysis of the effectiveness of lockdown and vaccination, the value of seven-day incidence is taken as the main characteristic of the intensity of the spread of the epidemic. The seven-day incidence is the difference in the intensity of infection over a seven-day period per 100,000 inhabitants of the age group under consideration.

Fig. 3 and Fig. 4 shows a comparison of the calculated and observed incidence values for the age groups 0-9 and 10-19, respectively. These graphs also show the incidence curve for Berlin. The original statistics are taken from [14].

**Figure 3.**
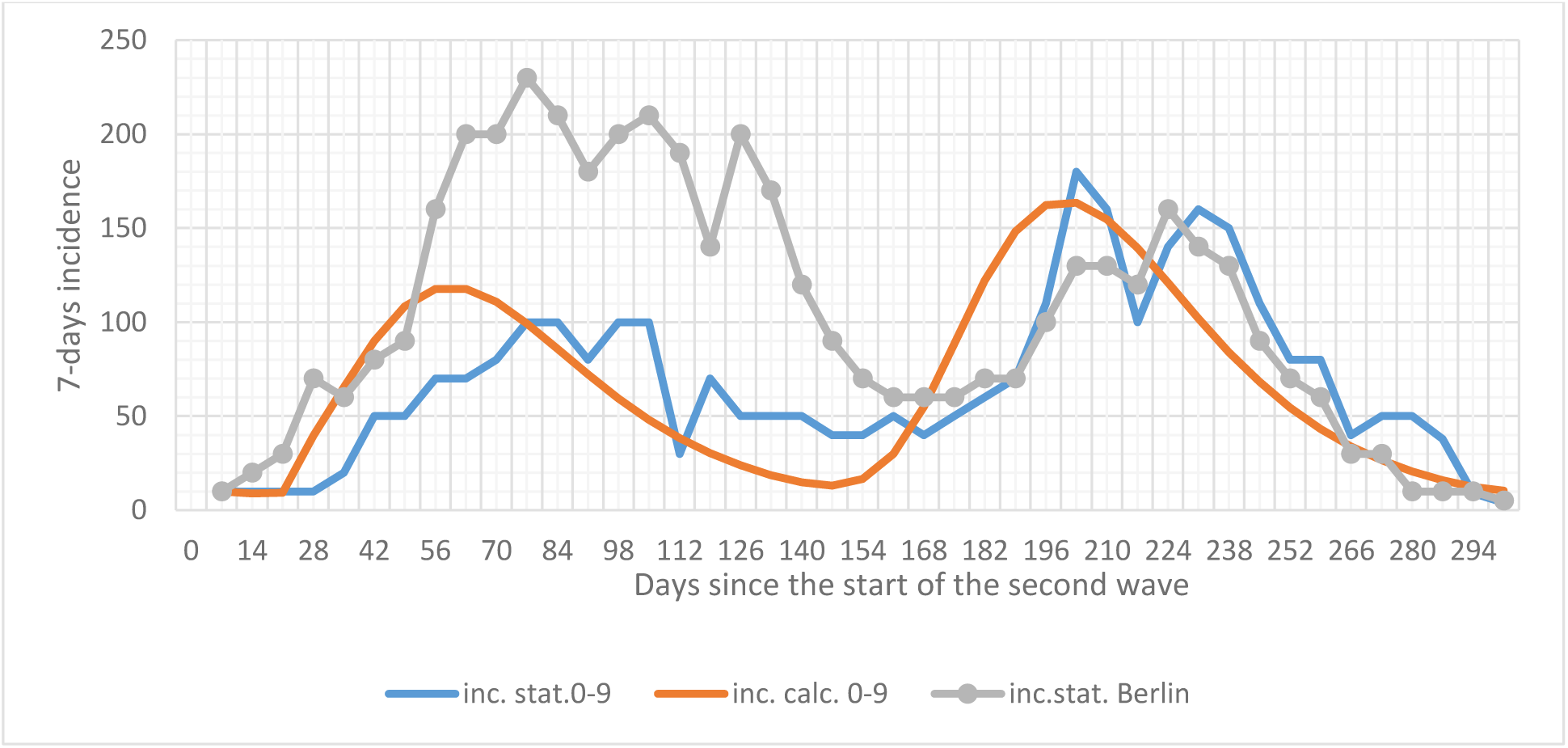
Seven-day incidence for the age group 0-9 years

**Figure 4.**
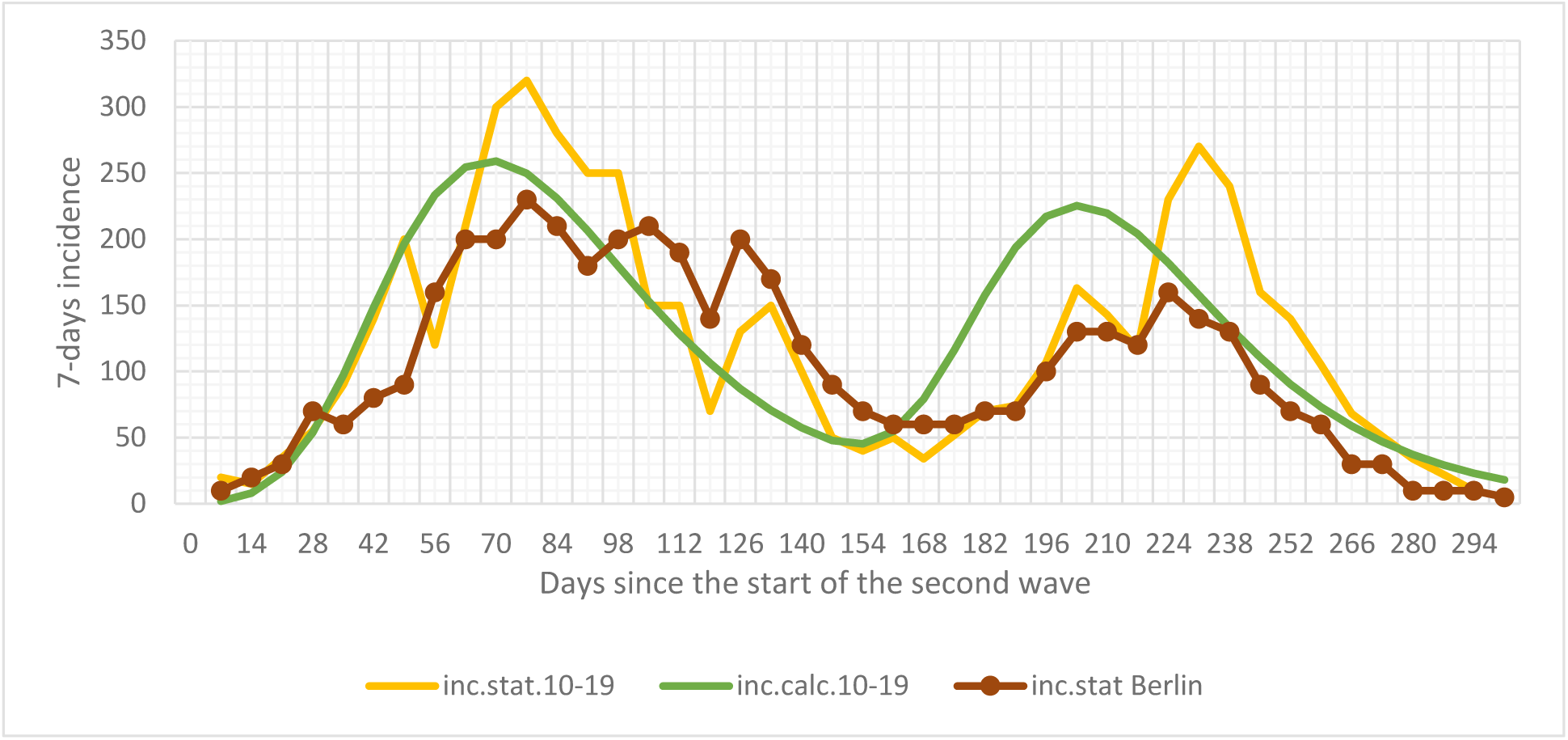
Seven-day incidence for the 10-19 age group

In these and subsequent graphs, the following notations are introduced:

inc.stat. are the incidence values for the indicated age groups and for Berlin according to observations, inc.calc. - calculated incidence data for the indicated age groups.

In general, the calculated and statistical data on incidence values are in good agreement qualitatively and quantitatively. As in Figure 2, there is a sharp divergence in the data for the different age groups. The intensity of the epidemic for the 0-9 age group is significantly lower than for the 10-19 age group and for Berlin in general. This is due both to the reduced level of contacts for children in this age group and probably also to their weaker susceptibility to the virus [18].

In the course of the present work, calculations were made for all ten age groups. In general, for each of the selected groups the calculation results agreed satisfactorily with the observational data, as shown in Figs. 2, 3 and 4.

The other age groups that are important to analyse are the elderly groups of 80-89 and 90+ year olds, which are relatively few in number and don’t determine the intensity of the epidemic in Berlin (the number of infections in these groups does not exceed 7% of the total number of infections in the city). But the number of infections in these groups largely determines the mortality rate from COVID-19.

Figure 6 compares the results of the calculations with the observed data for these age groups. The intensity of the epidemic in the 80-89 age group does not differ significantly from the average for Berlin. On the contrary, the infection rate in the 90+ age group is more than twice the average (the coefficient λ for the calculations was taken to be 0.0315 1/day). This is partly due to the fact that most residents of this age group live in old people’s homes, where diseases can become massive. In addition, the intensity of transmission for residents of this age group is also higher than the average for the general population.

**Fig. 6.**
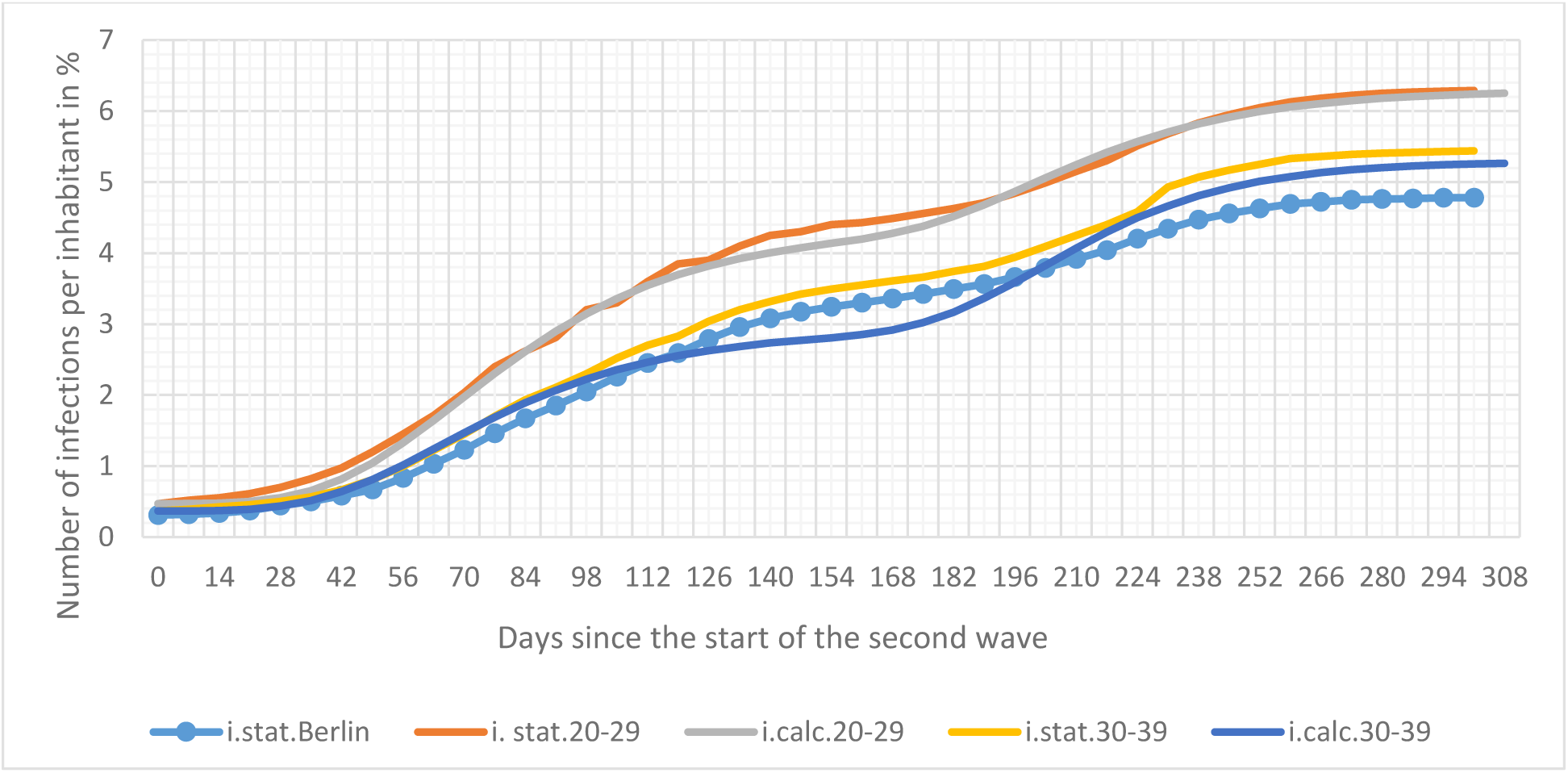
Spread of the epidemic in the age groups 80-89 and 90+

However, these two age groups are given priority for vaccination, so that by July of this year the percentage of people in these age groups was already over 80% vaccinated. As a result, since around 200 days after the start of the epidemic wave, there has been a steep decline in the number of infected people, as can be seen in Fig. 7 and especially Fig. 8.

**Figure 7.**
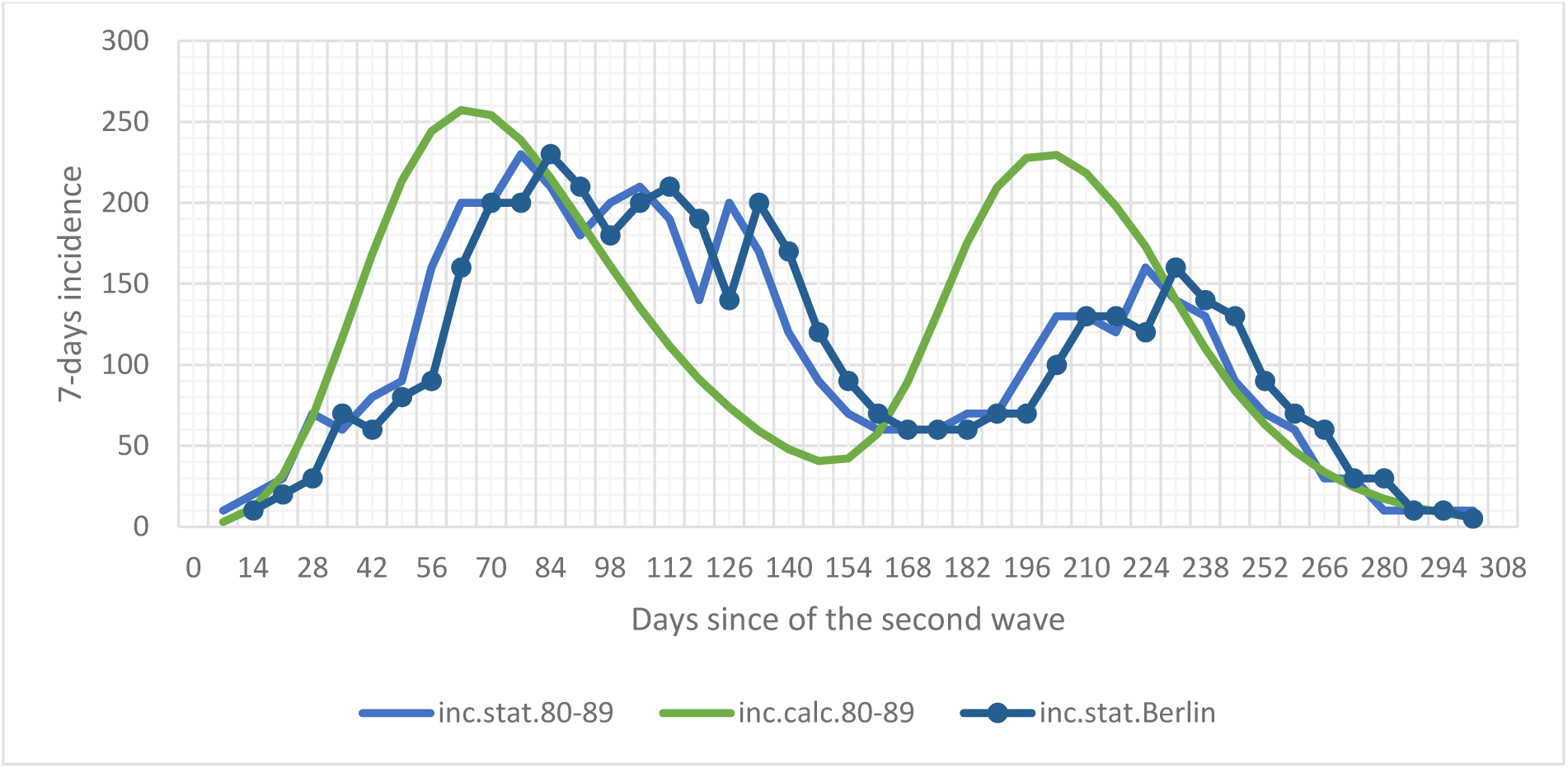
Seven-day incidence for the 80-89 age group

**Figure 8.**
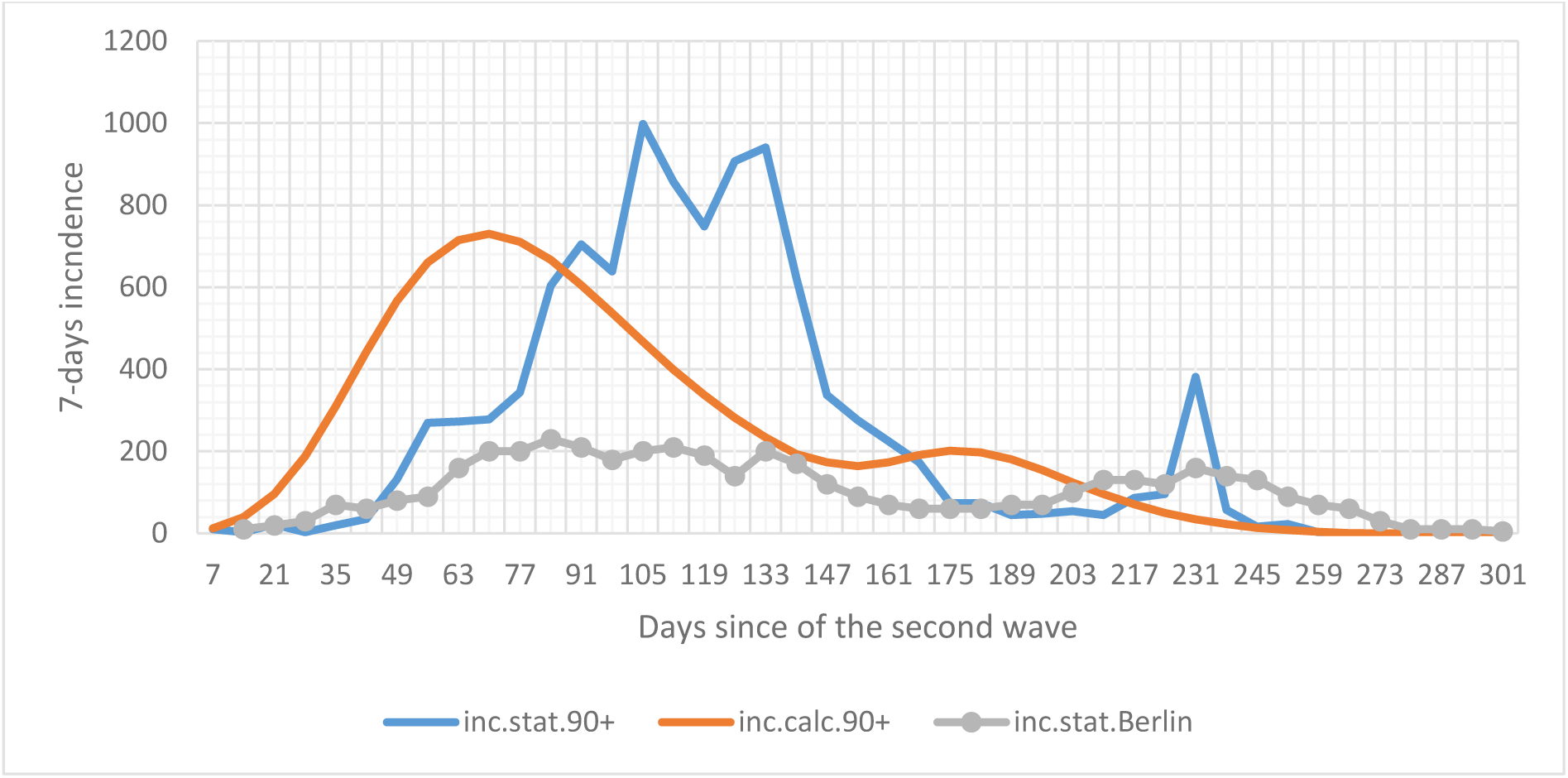
Seven-day incidence for the 90+ age group

Analysis of the graph in figure 8 shows that before vaccination the intensity of infection in the 90+ group was about 5 times higher than the average for Berlin. It was only as a result of mass vaccination that the virus epidemic in this age group was extinguished within a month.

The results of the calculation of the incidence rate for the 90+ age group differ significantly from the observed data, which is primarily due to the specific transmission patterns in this group, as mentioned above.

Overall, analysing the results of the calculations and comparing them with observational data, it can be generally concluded that the proposed ASILV model calculation methodology can be recommended for predicting the spread of the virus under conditions of lockdown and mass vaccination for individual age groups.

## Discussion

Main conclusion of the results was that comparing the calculated and statistical data on the spread of the epidemic in selected age groups shows that the ASILV model adequately describes the transmission patterns. The model calculations can therefore be used to analyse different situations involving the emergence of new, more transmissible strains of the virus, such as the delta variant, which could lead to new epidemic waves. The intensity of the new wave depends on the level of vaccination and the severity of the lockdown. As noted earlier, young people in their 20s and 40s have the most contacts and are minimally vaccinated. Figure 9 shows the size of each age group in relation to the total city population and the number of infected in the group in relation to the total number of infected.

**Fig. 9.**
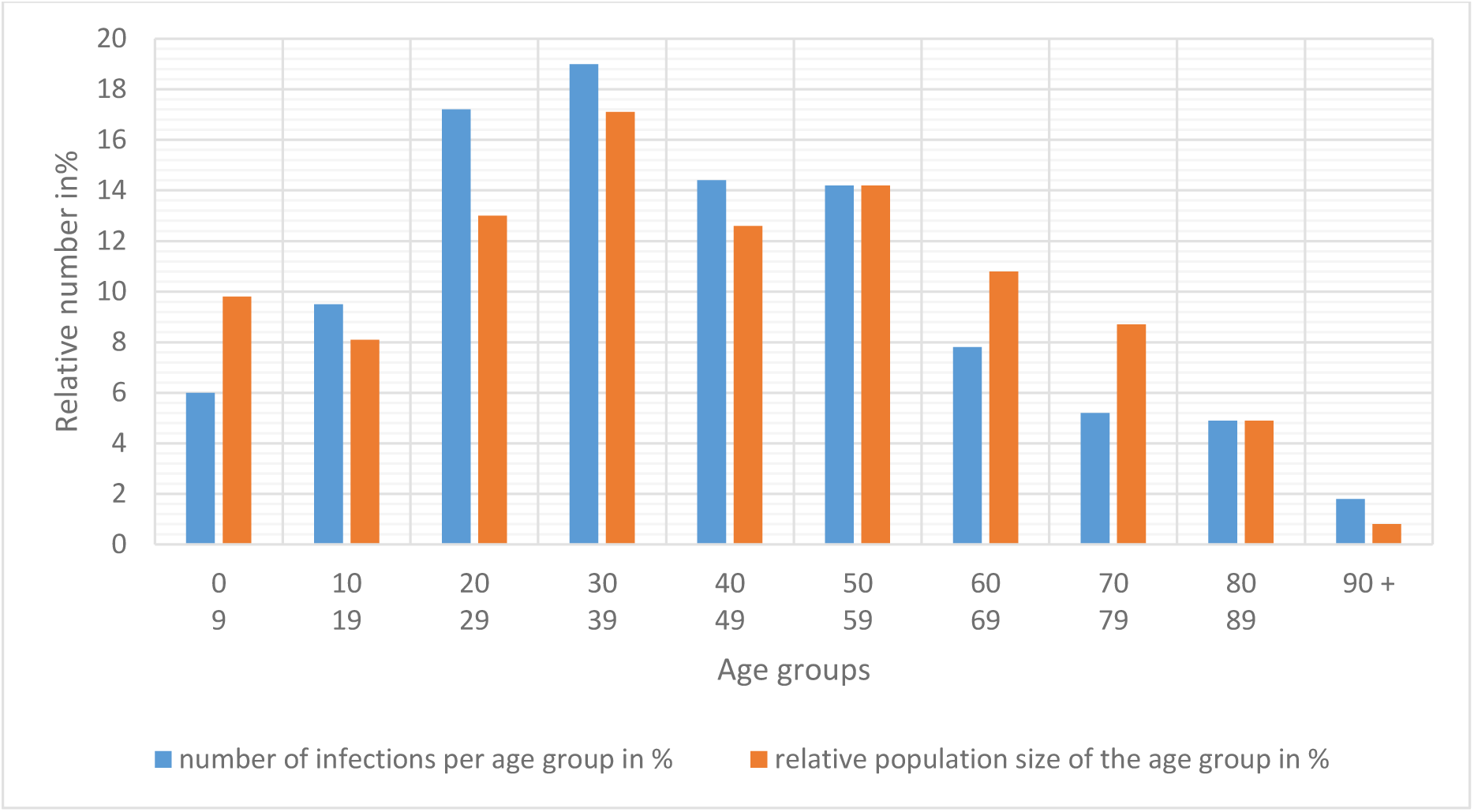
Relative number of infected persons for selected age groups.

As can be seen from figure 9, the relative number of infected persons in the 20-29 age group is about 30% higher than the relative number of persons in this group. Individuals in these age groups should be vaccinated first. In the case of school-age children in the 10-19 age group, their vaccination should be linked to the conditions for the beginning of the school season. In any case, given the possibility of mass development of the more transmissible delta variant in the city, relaxation of the lockdown before mass vaccination of 19- to 60-year-olds should be carefully considered. Variations in the relative number of infections for different age groups are closely related to variations in the coefficient λ of the estimated model. In turn, this coefficient, as noted earlier (see Fig. 1), depends on the level of lockdown efficiency for a particular group.

The values of the coefficients used in the model in the calculations for the different age groups are presented in Figure 10. It can be easily seen that the coefficient λ decreases as the ratio of the relative number of infected persons in an age group to its relative size increases. For example, the number of infected persons in the 90+ age group exceeds the size of this group relative to the total city population by a factor of 2.25, and the corresponding estimated coefficient for this group is minimal and equals λ = 0.0315 1/day.

**Figure 10.**
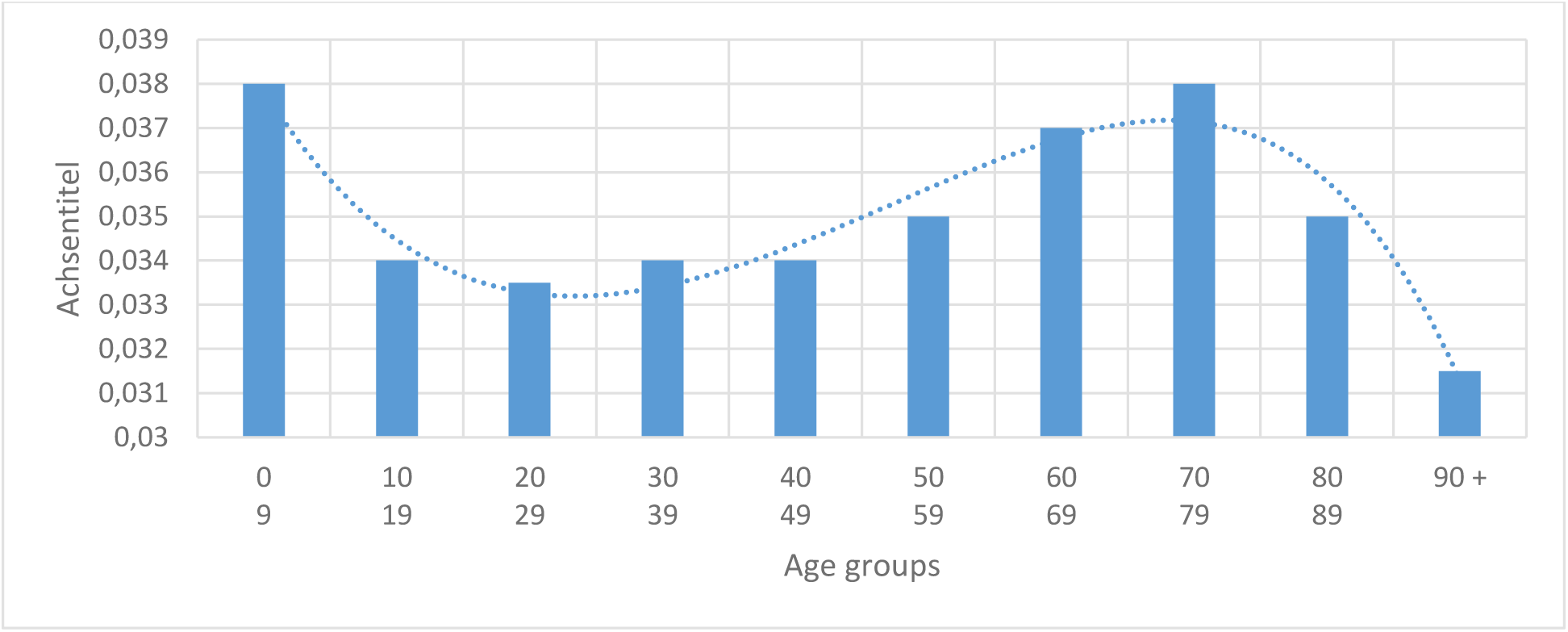
Values of coefficients λ for different age groups

The infection spread curves for all age groups show that, from about 300 days after the beginning of the second wave of the epidemic (at the beginning of July 2021), the seven-day incidence rate does not exceed 20 people per 100,000 inhabitants, with the highest incident rates at ages 10-19, 20-29, 30-39 and 40-49 years old where minimal vaccination has taken place. During the same period of time, the so-called alpha variant of the virus is most actively replaced by the more transmissible delta variant [19]. Vaccines against this type of virus are also 10-15% less effective than for the alpha variant [19]. As a consequence, a new rise in infection has been observed in some countries, such as the United Kingdom, Israel and the United States, where the epidemic has been virtually eliminated. Part of this new upsurge is due to a weakening of the lockdown conditions in these countries. An analysis of the data presented here shows that there is a high probability of a new upsurge in the epidemic in the 10-40 age groups, also in Berlin (at the time of writing, 1 July 2021).

We carry out control model calculations for the spread of the delta variant of the virus in unvaccinated age groups, which is characterised by increased transmission, as calculated by relation (3) by an increase of the K coefficient by 0.015 1/day compared to the alpha variant. Compared with the original variant of the virus this coefficient increases by 0.03 1/day.

The calculations were performed for two lockdown regimes: for typical conditions at λ = 0.035 1/day, when the intensity of the epidemic is reduced by a factor of 5 as a result of lockdown, and at λ = 0.033 1/day, that is, for a softer lockdown regime. Figure 11 presents the results of calculations of the seven-day incident rate at λ = 0.035 1/day, assuming that vaccination starts on a conditional zero-day, at the same time as the new epidemic wave (conditional beginning of July 2021). Given that by this time the vaccination rate had reached 30%, the calculated values presented in the figure must be divided by a factor of 1.5 in order to estimate the actual incidence rate for Berlin. The most realistic scenario for the new wave would be to raise the seven-day incidence rate to around 200 infected per 100,000 people in Berlin within roughly two months of the beginning of the new wave.

**Figure 11.**
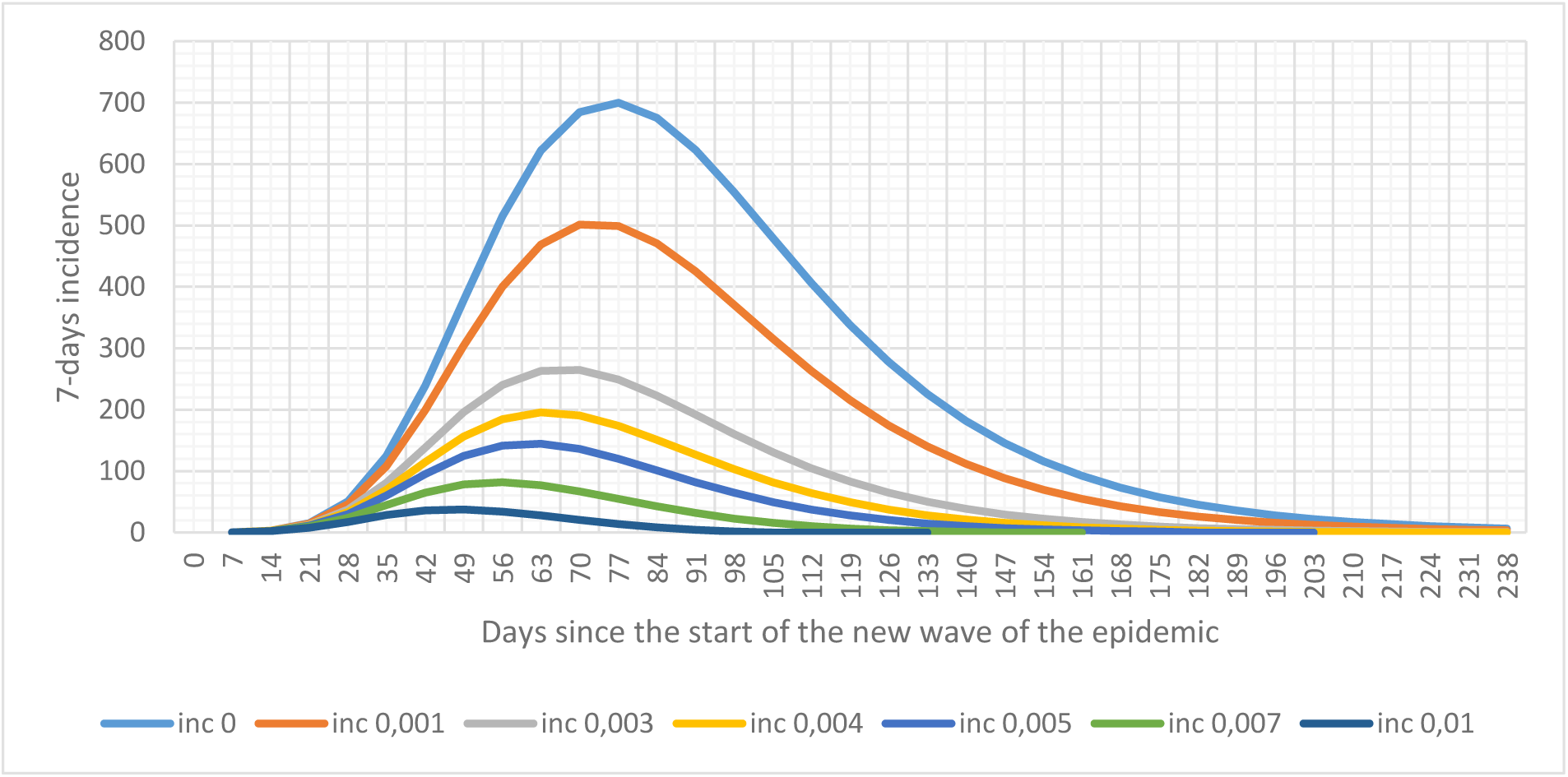
The value of the seven-day incidence for the delta variant at λ = 0.035. (Vaccination rates in brackets are 1/day.)

In an epidemic associated with exposure to the delta variant of the virus, there is an increased risk of infection in older persons who have received a full dose of vaccination. This is because in the case of the delta variant, the effectiveness of vaccination is reduced to 70-80% or even lower [19], i.e. there is a risk of illness in vaccinated inhabitants, but the probability of no vaccinated individuals being infected is about 3-4 times higher. However, it is important to emphasise that the likelihood of hospitalisation of vaccinated individuals remains very low.

If the lockdown conditions are relaxed, the intensity of the new epidemic wave increases.

Figure 12 shows the calculation of the value of the incidence when the coefficient λ = 0.033 1/day, or the value of the decrease in epidemic intensity by only a factor of 2. In this scenario, the maximum incidence increases to values of 250-300 infected.

**Fig. 12.**
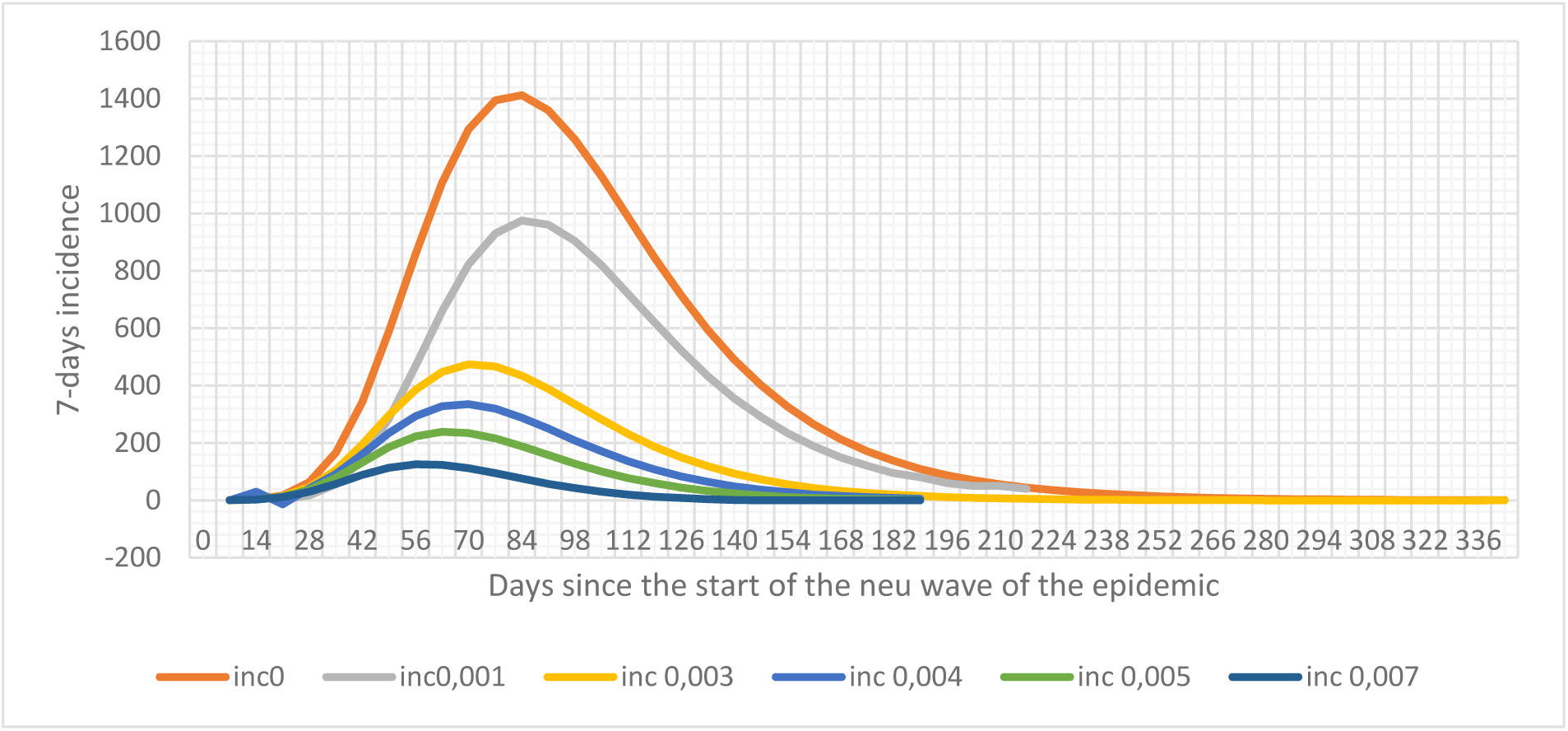
Seven-day incident size for the delta variant at λ = 0.033.

Thus, comparison of Fig.11 and Fig.12 suggests that further growth of the epidemic is expected if the lockdown limits are relaxed, with the most unfavourable scenario being the additional emergence of new, even more transmissible variants of the virus. Mass vaccination of residents in the 15 to 50 age groups could also prevent this scenario. With vaccination intensities of 0.005 1/day and above, the maximum incidence rate for calculated conditions does not exceed 100 per 100,000 inhabitants.

The sequelae of COVID 19 are closely related to the age of the patients. For age groups less than 40 years old no fatal cases of the virus have been reported in Berlin. The maximum number of fatal cases was observed for the 90+ age group [14]. Fig. 13 shows the ratio of fatal cases to the number of infected patients in the different age groups (as of 01 July 2021).

**Figure 13:**
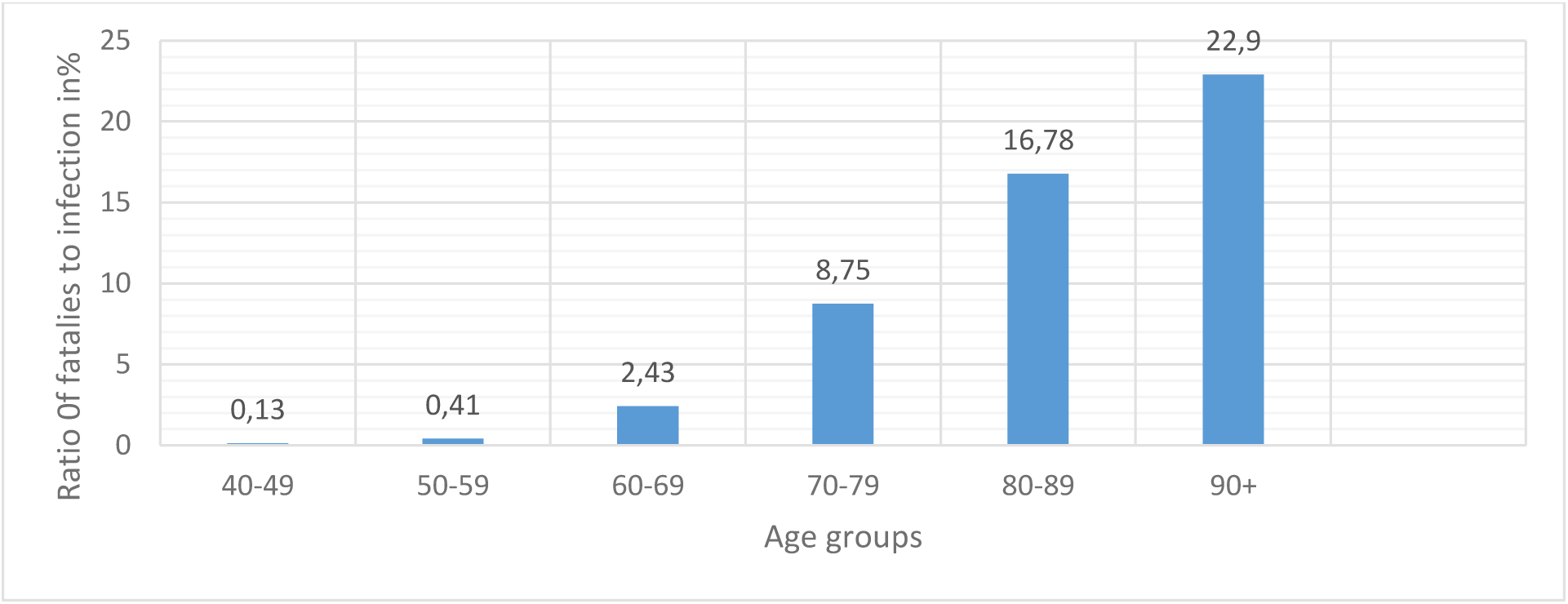
Ratio of fatalities to infections in % for different age groups.

The ordinate axis shows the percentage of fatalities divided by the total number of infected persons for the specified age groups.

Nearly one in six patients in the 80-89 age group and nearly one in four in the 90+ age group had a very high probability of dying before receiving vaccination.

Figure 14 shows how the number of fatalities from COVID-19 varies in these age groups in relation to increasing incidence. The abscissa axis shows the percentage of infected patients in the 80-89 and 90+ age groups, while the ordinate axis shows the percentage of fatalities in the total population in these age groups. The graph shows that the relative percentage of fatal cases increases with the number of infected individuals, reaching its maximum values, which are shown in Fig. 13.

**Figure 14:**
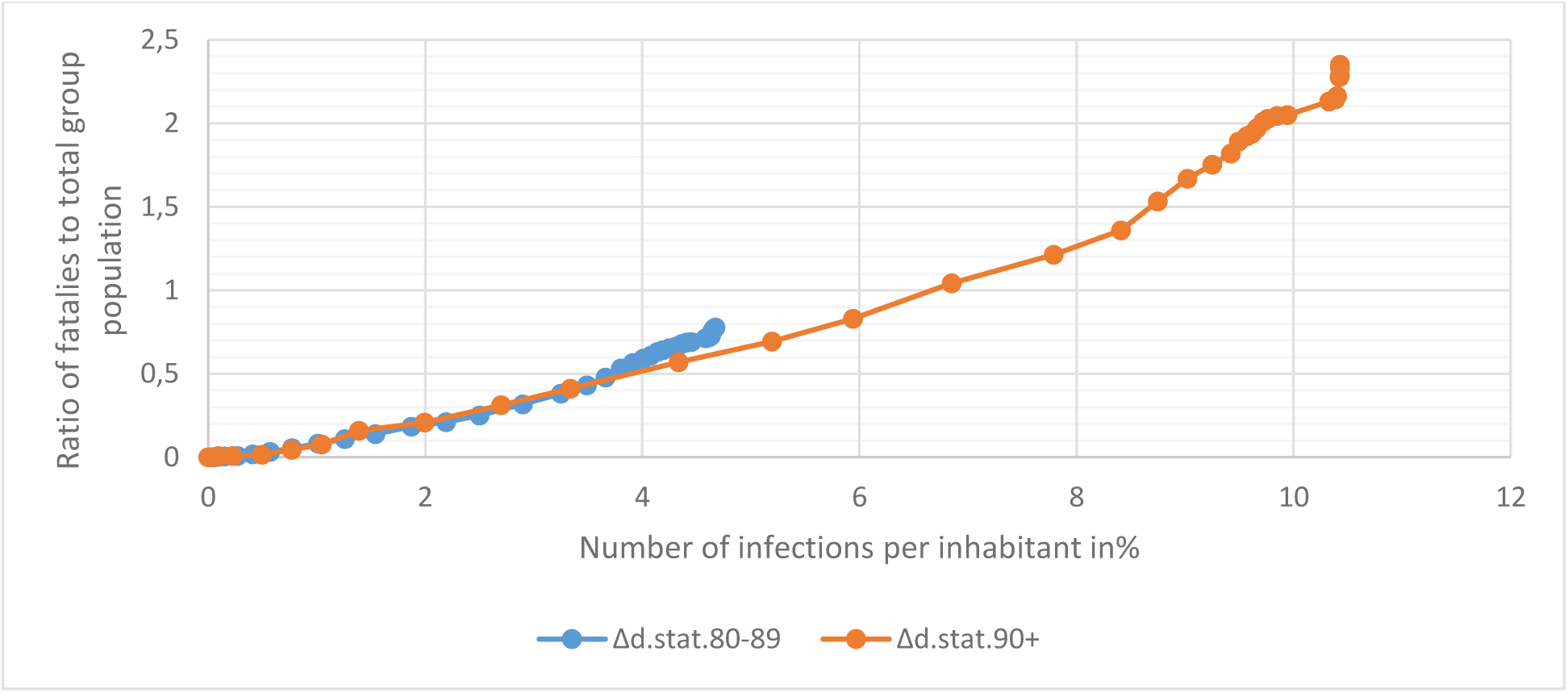
Ratio of fatalities to total population age groups 80-89 and 90+.

The data obtained make it possible to estimate the number of deaths for each age group d as a function of the relative number of infections i. These approximate ratios can be written down as follows:

for the 90+ group: d = 0.22 *i,

for the 80-89 group: d = 0.16 *i,

for the 70-79 group: d = 0.1 *i,

for the 60-69 group: d = 0.02 *i.

These ratios are valid for Berlin, and for other conditions should be adjusted if necessary. Importantly, the ASILV model can be used to estimate the number of infected patients in each age group, which in turn allows an approximate prediction of the likely number of fatalities associated with COVID-19.

## Conclusions

1. The previously developed ASILV model for calculating epidemic spread is modified to analyse the intensity of infection growth in selected age groups.
2. Comparison of the calculated spread and seven-day incidence values with corresponding observation data showed good agreement for the individual age groups.
3. Infection in the 20-40 age groups had the greatest impact on the overall spread of the epidemic. Relatively low vaccination rates at these ages induce new epidemic waves, particularly active when the virus mutates and isolation conditions become weaker.
4. The intensity of the epidemic in the 90+ age group has some features in comparison with other groups, which may be explained by differences in contact patterns among individuals in this age group.
5. Approximate ratios for estimating mortality rates as a function of infection intensity for individual age groups are given.
6. The proposed stratified ASILV model by age group will allow more detailed and accurate prediction of the spread of the COVID-19 epidemic, including when new, more transmissible versions of the virus mutate and emerge.

## Data Availability

statement regarding the availability of all data referred to in the manuscript

https://www.facebook.com/felix.mairanowski

